# Changes in Solo and Partnered Sexual Behaviors during the COVID-19 Pandemic: Findings from a U.S. Probability Survey

**DOI:** 10.1101/2020.06.09.20125609

**Authors:** Devon J. Hensel, Molly Rosenberg, Maya Luetke, Tsung-chieh Fu, Debby Herbenick

## Abstract

**Background:** Research demonstrates that pandemics adversely impact sexual and reproductive health (SRH), but few have examined their impact on people’s participation in sex. We examined self-reported changes in solo and sexual behaviors in U.S. adults during early stages of the public health response to COVID-19.

**Methods:** We conducted an online, nationally representative, cross-sectional survey of U.S. adults (N=1010; aged 18-94 years; 62% response rate) from April 10-20, 2020. We used weighted multinomial logistic regression to examine past month self-reported changes (decreased, stable or increased) in ten solo and partnered sexual behaviors. Predictor variables included: having children at home, past month depressive symptoms, (ACHA 3-item scale), past month loneliness (UCLA 3-Item Loneliness scale), COVID-19 protection behaviors (adapted 12-item scale), perceived COVID-19 consequences (adapted 10-item scale) and COVID-19 knowledge (adapted 10-item scale).

**Results:** Nearly half of all adults reported some kind of change – most commonly, a decrease – in their sexual behavior in the past month. Having elementary aged children at home, past month depressive symptoms and loneliness and enacting more COVID-19 protective behaviors were associated with both *reduced* partnered bonding behaviors, such as hugging, cuddling, holding hands and kissing, as well as *reduced* partnered sexual behaviors, such as oral sex, partnered genital touching and vaginal sex. Greater COVID-19 risk perception and greater COVID-19 knowledge were associated with mixed effects in behavior outcomes.

**Conclusions:** Our data illustrate the very personal ways in which different pandemic-associated factors may create or inhibit opportunities for solo and partnered sex. The centrality of sexuality to health and well-being – even during pandemics – means that a critical piece of public health prevention and management responses should is ensuring that services and resource that support positive sexual decision making remain open and available.

## Background

In March 2020, The World Health Organization^1^ (WHO) classified the novel coronavirus (SARS-CoV-2), and the illness it causes (COVID-19), as a pandemic. Past research demonstrates that pandemics (e.g., MERS, SARS, Ebola, Zika) adversely impact sexual and reproductive health (SRH), by reducing access to SRH-related supplies (e.g. contraception/condoms) or services (e.g. abortion, health care), by increasing exposure to sexual and/or physical violence, and by increasing economic insecurity.^2^ Few studies have examined how pandemics, including COVID-19, impact solo and partnered sexual behaviors.^3^ COVID-19 experiences could influence desire and ability to participate in sex, the types of sex they choose, and the extent to which behaviors differ from non-pandemic times. Understanding how and why changes occur is necessary to continue to adapt public health COVID-19 management in ways that are consistent with people’s fundamental rights to sexual health and well-being.^4,5^

COVID-19 transmission could influence how people approach sex. For example, COVID-19 can spread through aerosolized respiratory particles (e.g. coughs, sneezes, speaking). While preliminary data seem to indicate that the virus is detectable in some (e.g. semen and feces) but not all (e.g. vaginal fluid and urine) of the bodily fluids associated with sexual activities,^6^ more research is needed to understand sexual transmission risk. An infected person could transmit SARS-CoV-2 to a partner via airborne respiratory secretions or from their skin during close contact (e.g. kissing, genital touching), oral sex, vaginal sex, and/or sharing sex toys.^6,7^ Engaging in sexual behaviors that avoid close contact, like watching sexually explicit videos, sending sexual text messages, or phone/video sex, could reduce risk.^8^

What an individual knows about COVID-19 and how susceptible they feel to infection may also influence their sexual behavior. Individuals may avoid partnered, close contact sexual behaviors if they believe that getting COVID-19 may result in serious medical consequences for them^9^ or that the virus is difficult to treat.^10^ Individuals who are well-educated about COVID-19,^9^ including transmission and prevention information, may feel empowered to participate in partnered sex particularly if they perceive that knowledge makes them “low risk.“

Finally, COVID-19 mitigation measures, such as social distancing and recommended hygiene (e.g., hand washing), could impact how people approach sex. Many states and communities implemented “stay at home” orders that limit or prohibit activity in local non-essential businesses (e.g. shops, bars) and leisure areas (e.g. playgrounds) and have closed educational institutions (e.g. primary school and/or universities). Such policies have been associated with reduced sexual activity in men who have sex with men^11^ and Chinese adults.^12^ Moreover, teleworking, caring for children without childcare, school, or playdates/sleepovers, and helping school aged children engage in school online^13^ may impact both the time and space that people have available for sex. Social distancing may also exacerbate depression and loneliness,^14^ further decreasing desire for and frequency of sex.^15^ Meanwhile, social distancing or partner separation could increase solo masturbation or adjust partnered activities toward technology-mediated sexual interactions (e.g., using text messaging or video chat).^3,6,16^

The purpose of the current paper is to characterize the past month self-reported sexual changes in solo and partnered sexual behaviors in a nationally representative sample of U.S. adults, and understand how those changes are associated with structural, mental health, and COVID-associated risk perception and knowledge.

## Methods

### Study Design and Participants

Data were the 2020 National Survey of Sexual and Reproductive Health during COVID-19 (NSRHDC), a cross-sectional, online, nationally representative survey of COVID-19 related attitudes, experiences and knowledge among U.S. adults aged 18-94 years. The study was conducted in April 2020 by Ipsos Research using their KnowledgePanel® (Menlo Park, California) to recruit a probability-based web panel designed to be representative of noninstitutionalized U.S. citizens. Ipsos creates research panels using an address-based sampling (ABS) frame from using the U.S. Postal Service’s Delivery Sequence File – a database with full coverage of all delivery points in the U.S. ABS not only improves population coverage, but also provides a more effective means for recruiting hard-to-reach individuals, such as young adults, those without landline telephones and racial/ethnic minorities. Panel member households without internet connection are provided with a web-enabled device and free Internet service to maximize the breadth of participation.

The 26-item online survey took a median of 13 minutes (mode of 9 minutes) to complete, was available in English and Spanish languages, and was open for participation from April 10-20, 2020. Individuals randomly selected to participate were notified of the survey’s availability via email and through their online member page. Electronic notification allows surveys to be fielded more quickly and at less cost, and it also reduces the burden on participants, because they receive information in a more private fashion, and can take the survey at a time, and in a location that is convenient for them. KnowledgePanel® members typically only receive up to one survey per week, with an average of two to three per month. Reminder emails were sent to survey non-responders on the third day of the field period. Of the original individuals recruited (N=1632), 1010 (62%) completed the survey and represent the analytical sample in this study. Ipsos operates a modest incentive program that offers points for survey completion; points can be accumulated and exchanged for cash or merchandise.

Ipsos provided post-stratification, study-specific weights to adjust for any over- or under-sampling as well as nonresponse. Geodemographic distributions for the corresponding population were obtained from the CPS, the U.S. Census Bureau’s American Community Survey (ACS), or from the weighted KnowledgePanel profile data. For this purpose, an iterative proportional fitting procedure was used to produce the final weights. In the final step, calculated weights were examined to identify and, if necessary, trim outliers at the extreme upper and lower tails of the weight distribution. The resulting weights were then scaled to aggregate to the total sample size of all eligible respondents. Participant characteristics are included in Table 1.

**Table 1.** Unweighted and Weighted Participant Characteristics (N=1010)

| Characteristic | Unweighted | Weighted |
| --- | --- | --- |
|  | N (%) or Mean (SD) |  |
| Gender (female) | 516 (51.1) | 521 (51.6) |
| Sexual identity (minority: yes) | 68 (6.7) | 55 (5.9) |
| Gay or lesbian | 30 (3.1) | 31 (3.2) |
| Heterosexual | 896 (92.9) | 879 (91.9) |
| Bisexual | 27 (2.8) | 33 (3.4) |
| Something else | 11 (1.1) | 13 (1.4) |
| Age group (number of years) | 34.4 (17.1) | 29.8 (17.77) |
| 18-19 | 15 (1.5) | 27 (2.7) |
| 20-29 | 114 (11.3) | 184 (18.2) |
| 30-39 | 34 (3.3) | 175 (17.4) |
| 40-49 | 138 (13.7) | 142 (14.1) |
| 50-59 | 215 (21.3) | 185 (18.3) |
| 60-69 | 206 (20.4) | 165 (16.3) |
| 70-79 | 129 (12.8) | 103 (10.2) |
| 80+ | 37 (3.7) | 28 (2.8) |
| Household size (total number of people) | 2.65 (1.50) | 2.77 (1.60) |
| Any children less than five years (yes) | 113 (11.2) | 136 (13.4) |
| Any children less than 12 years (yes) | 127 (12.6) | 139 (13.8) |
| Any children 12-18 years (yes) | 105 (10.4) | 120 (11.9) |
| Current employment status |  |  |
| Working - as essential employee | 325 (33.6) | 336 (33.3) |
| Working - not as essential employee | 258 (26.7) | 253 (26.4) |
| Not working | 385 (38.1) | 368 (38.4) |
| Race/ethnicity (minority: yes) | 289 (28.6) | 371 (36.8) |
| White, non-Hispanic | 721 (71.4) | 639 (63.2) |
| Black, non-Hispanic | 85 (8.4) | 119 (11.8) |
| Other or 2+ races, non-Hispanic | 81 (8.0) | 87 (8.6) |
| Hispanic | 123 (12.2) | 165 (16.4) |
| Marital status |  |  |
| Married and/or living with partner | 668 (66.1) | 622 (61.6) |
| Separated/divorced | 129 (12.8) | 124 (12.3) |
| Widowed | 44 (4.4) | 40 (3.6) |
| Never married | 169 (16.7) | 224 (22.2) |
| Region |  |  |
| Northeast | 189 (18.7) | 177 (17.5) |
| Midwest | 228 (22.6) | 210 (20.8) |
| South | 341 (33.8) | 383 (37.9) |
| West | 252 (25.0) | 240 (23.8) |
| Household income |  |  |
| <\$25k | 95 (9.4) | 136 (13.5) |
| \$25-49k | 174 (17.2) | 184 (18.2) |
| \$50-74k | 183 (18.1) | 174 (17.2) |
| \$75-99k | 147 (14.6) | 141 (14.0) |
| \$100-124k | 108 (10.7) | 103 (10.3) |
| \$125-149k | 73 (7.2) | 66 (6.5) |
| \$150-174k | 90 (8.9) | 82 (8.1) |
| \$175k+ | 140 (13.9) | 124 (12.3) |
| Education |  |  |
| Less than high school | 75 (7.4) | 98 (9.7) |
| High school | 278 (27.5) | 295 (29.2) |
| Some college | 273 (27.0) | 281 (27.8) |
| Bachelor's degree or higher | 384 (38.0) | 336 (33.3) |
| Political orientation |  |  |
| Extremely liberal | 37 (3.9) | 87 (9.1) |
| Liberal | 149 (15.6) | 333 (35.0) |
| Slightly liberal | 85 (8.9) | 87 (9.1) |
| Moderate, middle of the road | 316 (33.0) | 333 (35.0) |
| Slightly conservative | 115 (12.0) | 113 (11.8) |
| Conservative | 209 (21.8) | 189 (19.9) |
| Extremely conservative | 47 (4.9) | 45 (4.7) |
| Depression symptoms (summed) | 1.54 (1.72) | 1.67 (1.78) |
| Lonely (yes) | 179 (17.7) | 190 (18.8) |

**Table 2.** Weighted Self-Reported Change Patterns in Solo and Partnered Sexual Behaviors during COVID-19 among U.S. Adults (N=1010).

| Pattern Type |  |
| --- | --- |
| Decrease only | 9.2 |
| Stable/no change only | 50.8 |
| Increase only | 5.4 |
| Increase and decrease | 3.1 |
| Increase and stable/no change | 11.9 |
| Decrease and stable/no change | 12.3 |
| Increase, decrease and stable/no change | 7.3 |

Ipsos provided post-stratification, study-specific weights to adjust estimates for any over- or under-sampling as well as nonresponse. Study procedures were approved by the Indiana University Institutional Review Board (#2004194314).

### Measures

#### Outcome variables

We assessed self-reported past month changes in ten solo and partnered sexual behavior categories: *hugged, kissed, held hands or cuddled with a romantic partner; masturbated by yourself; masturbated together with a partner or touched each other’s genitals; gave or received oral sex; engaged in penile-vaginal sex; sent or received sexy/nude pictures with a partner; watched erotica or porn with a partner; used a vibrator/sex toy during solo masturbation; used a vibrator/sex toy with a partner;* and *had a phone or video sex chat with a partner*. All items were originally assessed through a single prompt (“Since the new coronavirus started spreading in the US, to what extent have the following behaviors changed or stayed the same for you?”) using six response categories (much more, a little more, stable, a little less, much less, does not apply). We excluded “does not apply” and trichotomized the remaining data in each behavior (decreased [a little less and much less], stable, and increased [a little more and much more]) for analysis.

#### Predictor variables

The demographic predictor was c*hildren in the home* (all no/yes [referent]: any < 5 years, 6-12 year or 13+ years).

We also included two mental health variables. *Past month depressive symptoms* included a subset of five items (“Felt overwhelming anxiety,” “Felt very sad,” “Felt that things were hopeless,” “Felt so depressed that it was difficult to function” and “Felt very lonely” all (“No, never,” “Yes, but more than a month ago,” and “Yes, in the past month”) adapted from the American College Health Association.^17^ We created for analysis a dichotomous measure to indicate an answer of “Yes, in the past month” for one or more of the five items. *Loneliness* was examined using three items (“How often do you feel that you lack companionship,” “How often do you feel left out” and “How often do you feel isolated from others?” from the UCLA Loneliness Scale Short Form.^18^ We recoded each of the four original categories (“Not at all” [1], “Hardly ever” [1], “Some of the time” [3] and “Often” [4]) and summed all items. Finally, we created a dichotomous measure for analysis (not lonely [0-5] vs. lonely [6]).

We also included three COVID-19 context variables. *COVID-19 past month protection behaviors* was an index of 12, 4-point (not at all true of me, a little true of me, somewhat true of me, very true of me) behaviors (“I stayed at home,” “I did not physical attend social gatherings,” “I kept a distance of at least two meters (6 feet) from other people,” “If I had exhibited symptoms of sickness, I would have immediately informed the people around me,” “I washed my hands more frequently than the month before” and “I canceled out of state or international travel plans,” “I made sure I have enough prescription and over the counter medicines to meet my current health needs,” “I made sure I have enough condoms at home to meet my disease prevention and/or pregnancy prevention needs,” “I made sure I have enough birth control at home to meet my pregnancy prevention needs,” and “I made sure I had enough products to meet my hygiene needs [e.g. menstruation supplies, incontinence supplies, etc.”). The first six items were modified from COVID-19 research^19^ and the last six were designed by the second and last authors for this study.

### Perceived next month likelihood of COVID-19 consequences

was an additive index of ten, four point (High chance (> 75%), Medium chance (25-75%), Low chance (< 25%), No chance, Has already happened [dropped from analysis]) potential medical and social events (“being exposed,” “getting an infection,” “being hospitalized,” “knowing someone personally with an infection,” “knowing someone personally who has died of an infection,” “losing your job,” “partner/spouse losing their job,” “not having enough to eat,” “parent(s) losing their job,” “missing important healthcare appointments or treatment”). These were original items designed by the authors for this study.

### COVID-19 Knowledge

was an adapted^20,21^ ten-item scale designed to assess understanding of virus properties, transmission, and symptomology. A higher score indicates a greater depth of knowledge.

### Control variables

We included several control variables, including *gende*r, *race/ethnicity, sexual orientation* (heterosexual/sexual minority), *age, living with partner* (no/yes), *employment status* (working as an essential worker, working as non-essential worker, not working).

### Statistical Procedure

Mixed effects multinomial logistic regression (Stata, v.16) evaluated the impact of age of predictor variables on likelihood of reported change in each sexual behavior, controlling for the influence of confounding variables. In each model, we estimated the adjusted odds ratio (aOR) and corresponding 95% confidence interval (CI) for each predictor variable’s influence on an outcome behavior’s likelihood categories relative to the others. A random intercept approach allowed our estimates to vary across individuals, and adjusted estimates for clustering within states. We weighted all analyses to account for non-response and to adjust estimates to the demographic distribution in the United States. We evaluated each sexual behavior outcome twice - once using “stable” as the referent, and then using “decrease” as the referent (Tables 3 and 4).

**Table 3.** Weighted Self-Reported Changes in Solo and Partnered Sexual Behaviors during COVID-19 among U.S. Adults (N=1010).

| Solo and Partnered Sexual Behaviors | N | Decreased | No Change | Increased |
| --- | --- | --- | --- | --- |
|  |  | Percent |  |  |
| Hug, kiss, cuddle or hold hands with a romantic/sexual partner | 764 | 22.7 | 57.5 | 19.7 |
| Solo masturbation | 522 | 11.9 | 71.8 | 16.3 |
| Partnered masturbation or genital touching with a romantic/sexual partner | 439 | 12.8 | 76.7 | 10.5 |
| Gave and/or received oral sex | 506 | 18.1 | 71.7 | 10.2 |
| Penile-vaginal sex with a romantic/sexual partner | 577 | 19.4 | 68.0 | 12.5 |
| Send or receive sexy or nude pictures with a romantic/sexual partner | 290 | 8.9 | 80.1 | 11.0 |
| Watch sexually explicit videos (e.g. erotica or porn) | 385 | 12.0 | 72.2 | 15.8 |
| Use a vibrator or sex toy during solo masturbation | 296 | 13.2 | 81.9 | 4.8 |
| Use a vibrator or sex toy with a partner | 290 | 12.1 | 84.7 | 8.2 |
| Phone or video sex chat with a partner | 259 | 9.2 | 82.5 | 8.2 |

**Table 4.** Weighted adjusted multinomial odds ratios (aORs) – association of participant characteristics with reported part month changes in hugging, kissing, holding hands or cuddling with a partner, solo masturbation, partnered masturbation/genital touching and giving/receiving oral sex.

| Characteristics | Hugging, kissing, cuddling or holding hands with a partner |  |  | Solo masturbation |  |  |
| --- | --- | --- | --- | --- | --- | --- |
|  | Decrease vs. No Change (ref) | Increase vs. No change (ref) | Increase vs. Decrease (ref) | Decrease vs. No Change (ref) | Increase vs. No change (ref) | Increase vs. Decrease (ref) |
|  | aOR (95% CI)<sup>1</sup> |  |  | aOR (95% CI)<sup>1</sup> |  |  |
| Any children less than five years (yes) | **0.20 (0.09 - 0.45)\*\*\*** | 0.76 (0.40 - 1.47) | **3.70 (1.46 - 9.37)\*\*** | 0.44 (0.13 - 1.41) | 0.88 (0.39 - 2.30) | 1.98 (0.49 - 8.06) |
| Any children 5-12 years (yes) | **2.59 (1.39 - 4.82)\*\*** | 0.81 (0.40 - 1.62) | **0.31 (0.14 - 0.69)\*\*** | 1.21 (0.46 - 3.15) | 0.96 (0.34 - 2.71) | 0.78 (0.21 - 2.81) |
| Any children 13-18 years (yes) | 1.08 (0.56 - 2.06) | 1.22 (0.62 - 2.39) | 1.13 (0.51 - 2.50) | 1.51 (0.59 - 3.86) | 0.77 (0.26 - 2.25) | 0.52 (0.14 - 1.88) |
| Depressive symptoms past month (yes) | 1.15 (0.75 - 1.76) | 0.74 (0.47 - 1.15) | 0.64 (0.37 - 1.09) | 1.49 (0.76 - 2.89) | **4.28 (1.80 - 10.17)\*\*** | **2.86 (1.05 - 7.80)\*** |
| Lonely past month (yes) | **2.39 (1.50 - 3.80)\*\*\*** | 0.93 (0.51 - 1.69) | **0.39 (0.20 - 0.74)\*\*** | 0.67 (0.31 - 1.45) | 0.88 (0.43 - 1.80) | 1.31 (0.52 - 3.32) |
| Chance of COVID-related consequences | 1.03 (0.99 - 1.08) | 1.01 (0.97 - 1.06) | 0.97 (0.92 - 1.03) | **1.10 (1.03 - 1.16)\*\*** | 1.06 (0.99 - 1.13) | 0.96 (0.89 - 1.03) |
| Protective/preparation behaviors | **1.05 (1.01 - 1.09)\*** | 1.02 (0.98 - 1.07) | 0.97 (0.92 - 1.02) | **1.09 (1.03 - 1.16)\*\*** | 1.04 (0.98 - 1.11) | 0.95 (0.88 - 1.02) |
| COVID knowledge | 0.98 (0.85 - 1.14) | 0.88 (0.76 - 1.02) | 0.89 (0.75 - 1.07) | 0.83 (0.67 - 1.02) | **0.73 (0.59 - 0.90)\*\*** | 0.88 (0.68 - 1.13) |
| Characteristics | Partnered masturbation and/or genital touching |  |  | Giving and/or receiving oral sex |  |  |
|  | Decrease vs. No Change (ref) | Increase vs. No change (ref) | Increase vs. Decrease (ref) | Decrease vs. No Change (ref) | Increase vs. No change (ref) | Increase vs. Decrease (ref) |
|  | aOR (95% CI)<sup>1</sup> |  |  | aOR (95% CI)<sup>1</sup> |  |  |
| Any children less than five years (yes) | 0.75 (0.25 - 2.19) | **0.21 (0.05 - 0.86)\*** | 0.28 (0.05 - 1.50) | **0.13 (0.04 - 0.43)\*\*** | 0.39 (0.13 - 1.16) | 2.88 (0.66 - 12.50) |
| Any children 5-12 years (yes) | 0.93 (0.28 - 3.01) | **3.26 (1.18 - 8.98)\*** | 3.51 (0.82 - 14.94) | 2.11 (0.90 - 4.93) | 1.92 (0.72 - 5.07) | 0.91 (0.28 - 2.88) |
| Any children 13-18 years (yes) | 1.32 (0.47 - 3.73) | 0.44 (0.12 - 1.55) | 0.33 (0.07 - 1.52) | 0.99 (0.41 - 2.37) | 0.96 (0.32 - 2.82) | 0.96 (0.28 - 3.33) |
| Depressive symptoms past month (yes) | 1.17 (0.53 - 2.55) | 0.97 (0.42 - 2.21) | 0.82 (0.28 - 2.38) | **2.77 (1.48 - 5.39)\*\*** | 1.39 (0.62 - 3.07) | 0.50 (0.19 - 1.29) |
| Lonely past month (yes) | **3.66 (1.78 - 7.56)\*\*\*** | 0.74 (0.25 - 2.18) | **0.20 (0.06 - 0.66)\*\*** | **2.74 (1.48 - 5.07)\*\*** | 0.80 (0.29 - 2.15) | **0.29 (0.10 - 0.82)\*** |
| Chance of COVID-related consequences | 1.02 (0.95 - 1.09) | **1.09 (1.01 - 1.16)\*** | 1.06 (0.97 - 1.16) | 1.05 (0.99 - 1.11) | 1.02 (0.95 - 1.09) | 0.97 (0.89 - 1.05) |
| Protective/preparation behaviors | **1.11 (1.03 - 1.18)\*\*** | 1.05 (0.98 - 1.13) | 0.94 (0.86 - 1.03) | 1.05 (0.99 - 1.11) | 1.04 (0.97 - 1.12) | 0.99 (0.92 - 1.07) |
| COVID knowledge | 0.83 (0.66 - 1.04) | **0.71 (0.57 - 0.90)\*\*** | 0.86 (0.65 - 1.14) | **0.75 (0.62 - 0.91)\*\*** | **0.73 (0.58 - 0.91)\*\*** | 0.96 (0.75 - 1.24) |

<sup>1</sup>Controls are gender, race/ethnicity, sexual orientation, age, living with partner and work status
\* $p < .05$ ; \*\* $p < .01$ ; \*\*\* $p < .001$

## Results

### Participant characteristics

The weighted sample was 51.5% female, 28.6% ethnic/racial minority (8.4% non-Hispanic Black, 12.2% Hispanic, 8.2% other or multiple races) with a mean age of 34.4 years (SD=17.1 years; range: 18-94 years). Most were heterosexual (92.9%) and married and/or cohabitating (66.6%). About a third were employed as either an essential worker (33.6%). The mean household size was about two people (SD=1.5) (Table 1).

### Prevalence of changes in solo and partnered sexual behavior

Overall, across all ten behaviors (Tables 2 and 3), nearly half (49.2%) of the sample reported some kind of change – most commonly, a decrease – in their sexual behavior in the past month. The most common behavior (Table 2) to *increase* were hugging, kissing, cuddling or holding hands with a partner (19.7%) and the least common increase was vibrator or sex toy use during solo masturbation (9.2%). The most common behavior (Table 2) to *decrease* was hugging, kissing, cuddling or holding hands with a partner (22.7%) and the least common decrease was sending or receiving sexy or nude pictures from a partner (8.9%). Over half of participants (57.5%) reported stability in hugging, kissing, cuddling or holding hands with a partner. Most reported stability in sending or receiving sexy or nude pictures from a partner (80.1%), vibrator or sex toy use during solo masturbation (81.9%), having phone or video chat sex with a partner (82.5%) and vibrator or sex toy during partnered masturbation (84.7%)

### Multinomial Regression Results

We evaluated all outcomes (Tables 4 and 5) using both “no change” and “decreased” in turn as referent categories. Significant results from all models are reported below.

**Table 5.** Weighted adjusted multinomial odds ratios (aORs) – association of participant characteristics with reported part month changes in vaginal sex, sending/receiving sex or nude photos, watching porn/erotica with a partner or had phone/video sex or chat with a partner.

| Characteristics | Vaginal sex |  |  | Sent or received sexy or nude photos with partner |  |  |
| --- | --- | --- | --- | --- | --- | --- |
|  | Decrease vs. No Change (ref) | Increase vs. No change (ref) | Increase vs. Decrease (ref) | Decrease vs. No Change (ref) | Increase vs. No change (ref) | Increase vs. Decrease (ref) |
|  | aOR (95% CI)<sup>1</sup> |  |  | aOR (95% CI)<sup>1</sup> |  |  |
| Any children less than five years (yes) | **0.28 (0.12 - 0.66)\*\*** | 0.50 (0.21 - 1.21) | 1.80 (0.60 - 5.36) | 1.32 (0.31 - 5.53) | 0.55 (0.13 - 2.35) | 0.42 (0.06 - 2.84) |
| Any children 5-12 years (yes) | **3.13 (1.53 - 6.40)\*\*** | 1.76 (0.77 - 4.00) | 0.56 (0.21 - 1.42) | 0.24 (0.02 - 2.21) | 1.30 (0.29 - 5.80) | 5.44 (0.40 - 72.98) |
| Any children 13-18 years (yes) | 1.10 (0.53 - 2.28) | 1.17 (0.50 - 2.71) | 1.07 (0.40 - 2.82) | 1.04 (0.20 - 5.27) | 0.44 (0.07 - 2.58) | 0.40 (0.04 - 3.74) |
| Depressive symptoms past month (yes) | **2.36 (1.35 - 4.13)\*\*** | 0.83 (0.44 - 1.56) | **0.34 (0.16 - 0.75)\*\*** | 0.53 (0.15 - 1.77) | 0.88 (0.26 - 2.91) | 1.68 (0.34 - 8.37) |
| Lonely past month (yes) | **2.12 (1.20 - 3.71)\*\*** | 1.16 (0.52 - 2.58) | 0.55 (0.23 - 1.31) | 1.46 (0.44 - 4.80) | 1.49 (0.50 - 4.40) | 1.02 (0.23 - 4.46) |
| Chance of COVID-related consequences | 1.05 (0.99 - 1.10) | **1.08 (1.02 - 1.15)\*\*** | 1.03 (0.96 - 1.10) | **1.19 (1.08 - 1.30)\*\*\*** | **1.09 (1.00 - 1.20)\*** | 0.91 (0.81 - 1.02) |
| Protective/preparation behaviors | **1.07 (1.02 - 1.12)\*\*** | 1.02 (0.96 - 1.08) | 0.95 (0.89 - 1.02) | 1.01 (0.92 - 1.10) | 0.94 (0.87 - 1.02) | 0.93 (0.83 - 1.03) |
| COVID knowledge | 0.86 (0.72 - 1.03) | **0.80 (0.66 - 0.98)\*** | 0.93 (0.74 - 1.16) | **0.69 (0.51 - 0.93)\*** | 0.80 (0.60 - 1.06) | 1.15 (0.80 - 1.66) |
| Characteristics | Watched porn or erotica with a partner |  |  | Had phone or video sex/chat with a partner |  |  |
|  | Decrease vs. No Change (ref) | Increase vs. No change (ref) | Increase vs. Decrease (ref) | Decrease vs. No Change (ref) | Increase vs. No change (ref) | Increase vs. Decrease (ref) |
|  | aOR (95% CI)<sup>1</sup> |  |  | aOR (95% CI)<sup>1</sup> |  |  |
| Any children less than five years (yes) | 0.47 (0.13 - 1.62) | 0.47 (0.15 - 1.44) | 1.01 (0.21 - 4.86) | 0.33 (0.05 - 2.17) | 0.17 (0.01 - 1.99) | 0.55 (0.02 - 10.50) |
| Any children 5-12 years (yes) | 2.48 (0.79 - 7.75) | 2.52 (0.85 - 7.43) | 1.01 (0.24 - 4.30) | 1.67 (0.22 - 12.60) | 1.05 (0.12 - 9.06) | 0.68 (0.04 - 11.47) |
| Any children 13-18 years (yes) | 0.57 (0.15 - 2.20) | 0.66 (0.21 - 2.08) | 1.14 (0.22 - 5.95) | 0.89 (0.10 - 7.62) | 3.59 (0.60 - 21.23) | 4.18 (0.32 - 53.95) |
| Depressive symptoms past month (yes) | 0.96 (0.40 - 2.30) | 1.86 (0.77 - 4.47) | 1.93 (0.61 - 6.05) | 0.97 (0.18 - 5.29) | 3.10 (0.50 - 19.21) | 4.16 (0.36 - 47.97) |
| Lonely past month (yes) | 1.31 (0.53 - 3.21) | 0.96 (0.40 - 2.30) | 0.73 (0.23 - 2.30) | **5.63 (1.30 - 24.28)\*** | 1.41 (0.30 - 6.62) | 0.25 (0.03 - 1.75) |
| Chance of COVID-related consequences | **1.07 (1.00 - 1.15)\*** | 1.05 (0.98 - 1.12) | 0.97 (0.89 - 1.06) | **1.22 (1.08 - 1.39)\*\*** | 1.03 (0.90 - 1.17) | **0.82 (0.69 - 0.98)\*** |
| Protective/preparation behaviors | **1.10 (1.02 - 1.19)\*\*** | 0.97 (0.90 - 1.05) | **0.88 (0.80 - 0.97)\*** | 1.03 (0.92 - 1.15) | 1.01 (0.89 - 1.13) | 0.97 (0.84 - 1.13) |
| COVID knowledge | **0.76 (0.60 - 0.97)\*** | 0.78 (0.61 - 1.00) | 1.01 (0.75 - 1.38) | **0.58 (0.40 - 0.84)\*\*** | **0.49 (0.33 - 0.73)\*\*\*** | 0.87 (0.53 - 1.42) |

<sup>1</sup>Covariates are gender, race/ethnicity, sexual orientation, age, living with partner and work status
\* $p < .05$ ; \*\* $p < .01$ ; \*\*\* $p < .001$

### Children and sexual behavior

Participants with any children under age five in the house were three times *more likely* to report increased (vs. decreased: aOR=3.70) hugging, kissing, cuddling or holding hands with a partner in the past month, and were *less likely* to report a decrease (vs. stable: aOR [95% CI]=0.20 [0.09-0.45]) in the same behavior. Having children in this age range was linked to participants being less likely to report increased partner masturbation/genital touching (vs stable: aOR [95% CI]=0.21 [0.05-0.86]), as well as a reduced chance that oral or vaginal sex had decreased (vs. stable: aOR [95% CI]=0.13 [0.04-0.43]) and 0.28 [0.12-0.66]).

Having children 6-12 years old in the house was linked to a lower chance of reporting increased (vs. decreased: aOR [95% CI]=0.31 [0.14-0.69]) hugging, kissing, cuddling or holding hands with a partner in the past month. Having children in this age range was linked to a higher likelihood of reporting both that partnered masturbation/genital touching had increased (vs stable; aOR [95% CI]=3.26 [1.18-8.98]), and that reporting that vaginal sex had decreased (vs. stable; aOR [95% CI]=3.13 [1.53-6.40]) in the past month.

### Depression, loneliness and sexual behavior

Participants who reported one or more depressive symptoms in the past month were three to four times more likely to report increased solo masturbation (vs. stable: aOR [95% CI]=4.28 [1.80-10.17]) Depressive symptoms also made a decrease in past month oral sex and vaginal sex more likely (vs. stable: aOR [95% CI]=2.77 [1.48-5.39]) and 2.36 [1.35-4.13]). Past month loneliness was associated with a greater likelihood that hugging, kissing, cuddling or holding hands with a partner (aOR [95% CI]=2.39 [1.50-3.80]) and partnered masturbation/genital touching (aOR [95% CI]=3.66 [1.78-7.56]), any oral sex (aOR [95% CI]=2.74 [1.48-5.07) and vaginal sex (aOR [95% CI]=2.36 [1.35-4.13]) had decreased (vs. remained stable) in the past month.

### COVID-19 consequences perception and sexual behaviors

Participants with a greater belief in the likelihood of COVID-19 was linked to a higher likelihood of reporting that partnered masturbation (aOR [95% CI]=1.09 [1.01-1.16]), vaginal sex (aOR [95% CI]=1.08 [1.02-1.15]) and sending/receiving sexy or nude photos with a partner (aOR [95% CI]=1.09 [1.00-1.20]) had increased (vs. stable) in the past month. Greater consequence perception was also linked to a greater chance that participants reported decreased (vs. stable) solo masturbation (aOR [95% CI]=1.10 [1.03-1.16]) or watching porn/erotica with a partner (aOR [95% CI]=1.07 [1.01-1.15]) in the past month.

### COVID-19 protection behaviors and sexual behaviors

Enacting more protective behaviors was linked to decreased (vs. stable) in hugging, kissing, cuddling or holding hands with a partner, solo masturbation partnered masturbation/genital touching, vaginal sex and watching porn/erotica with a partner in the past month (all aOR=1.03-1.10).

### COVID-19 knowledge and sexual behaviors

Finally, greater COVID-19 knowledge was associated with lower likelihood of increased (vs. stable) solo masturbation, partnered masturbation/genital touching, receiving oral sex, vaginal sex and phone/video sex/chat with a partner in the past month (aOR=0.49-0.80 for all outcomes). Participants with greater COVID-19 knowledge were less likely to report they had experienced decreased (vs stable) giving/receiving oral sex, sending/receiving sexy or nude photos with a partner, watching porn/erotica with a partner and phone/video sex/chat with a partner in the past month (aOR=0.58-0.75 for all outcomes).

## Discussion

Past research demonstrates that pandemics (e.g., MERS, SARS, Ebola, Zika) adversely impact sexual and reproductive health (SRH),^2^ yet limited scientific evidence – including from the COVID-19 pandemic – examines their influence on how people organize their sexual lives. Our work is one of the first studies to address this literature gap, using a U.S. nationally representative, probability survey of adults to assess self-reported changes in solo and partnered sexual behaviors relatively early in the COVID-19 pandemic. Half of all adults in the United States reported that they had experienced change – most commonly, a decrease – in their past month sexual behavior (a time when most of the country was subject to stay at home guidance).

Our data illustrate the very personal ways in which different pandemic-associated factors may create or inhibit opportunities for solo and partnered sex. We found that having any children under the age of five at home was associated with *greater* likelihood of stability and/or increase in several partnered behaviors, while having elementary aged children was often linked to *decreased* reports of these behaviors. These findings are largely consistent with differences in how “stay at home” orders may have asked parents to balance working from home and childcare in an absence of school, day care, or other forms of childcare outside the home. Parents of smaller children may be better able to maintain pre-pandemic schedules and routines (e.g. naptimes and/or earlier bedtimes) that free some consistent time for partnered sex in ways that may be more challenging for parents of older school-aged children to do.^13^ Parents of young children may also report increased hugging, kissing, cuddling or holding hands because it is a part of group/family interactions (e.g., family cuddling, snuggling, or hugging) which may increase oxytocin and decrease cortisol, at least for mothers.^22^

Further, although we did not measure perceived domestic stress, it could also be that integrating multiple roles (e.g. parent, educator, etc.) may create stress that lowers desire either for sex or for extended time with one’s partner.^12,23^ Then again, people engage in solo and partnered sex for varied reasons including due to boredom and to relieve stress, and partnered sex is well-documented to enhance love, pleasure, and nurturance.^24-26^ Global research has raised concerns that social distancing measures, though necessary to control COVID-19, can exacerbate feelings of depression and loneliness for some people.^14^ We add to this body of research by showing that past month depressive symptoms and loneliness were associated with both *reduced* partnered bonding behaviors, such as hugging, cuddling, holding hands and kissing, as well as *reduced* partnered sexual behaviors, such as oral sex, partnered genital touching and vaginal sex.

Somewhat consistent with existing research,^9,10^ we found that people who perceived greater personal risk for COVID-19 medical or (e.g., they or a loved one getting sick) or social consequences (e.g. job loss or missing medical appointments) reported a decrease in some, but not all, solo and partnered sexual behaviors. Subsequent research might assess whether how specific sexual transmission concerns, as well as more global pandemic-associated stress, impact people’s sexual choices. Finally, our research shows that people with greater COVID-19 knowledge were more likely to report stability in partnered sexual behaviors. We could not assess whether greater knowledge increased people’s comfort to maintain existing habits, or whether greater knowledge was a barrier to more sex. Future studies that assess the link between coronavirus knowledge and subsequent sexual behaviors would benefit from including scale items that specifically address sex as a transmission route.

### Implications

Our data answer a call for research that more explicitly integrates sexuality and sexual behavior within the pandemic literature,^5^ but more research is needed in order to better understand the short- and long-term effects of the pandemic experience on human sexual health and well-being. A research agenda that maintains a focus on sexual well-being during pandemics supports existing public health prevention goals, as well as global perspectives on sexual and reproductive health rights.

From a *public health perspective*, sexual-health-as-prevention paradigms recognizes that an individual’s circumstances impact their decisions about how and when to have sex.^27-32^ Key resources like access to health care and services, access to condoms/contraception, and access to medications for sexual health, scaffold people’s ability to control when they want to have sex, make sex more enjoyable, and reduce and/or avoid risk behaviors when sex occurs. Pandemics cause well-documented interruptions to all of these resources,^2^ increasing people’s downstream exposure to adverse outcomes like sexual violence, unintended pregnancy and sexually transmitted infections, as well as the mental and sexual health consequences of unwanted abstinence.^33^ These effects may particularly be magnified in sexual and/or racial minority populations.^11^ As part of pandemic preparation, communities should ensure that sexual health care facilities remain open, all sexual and reproductive health care services remain “essential” and that supply chains of key supplies remain as open as possible.

From a *global sexual and reproductive health rights perspective*, many international health organizations have increasingly endorsed both the idea that sexuality is a central element in life long health and well-being^34,35^ and that access to experiences that promote positive sexuality – including sexual behavior – are a human right.^36^ In the context of pandemics, this perspective affirms the role that evidence-based research plays in helping clinicians and health educators understand the importance of sexuality and sexual experience during times of public crisis, assess individual barriers to desired sexual experiences, and devise solutions to sexual challenges that are appropriately tailored within a person’s circumstances. The near complete lack of sexual behavior research amidst the explosion of other COVID-19 health outcomes research may inadvertently lead health professionals and lay people alike to falsely conclude that individuals should universally refrain from sex with non-household partners until the pandemic ends. Messaging that encourages sexual abstinence or that stigmatizes sexual desire is unlikely to reduce sex, and in fact may compound people’s existing pandemic-associated mental health challenges and/or prevent 13 them from seeking risk reduction advice from their health care provider.^6^ Our research, as well as future pandemic sexual health research, could help prepare clinicians and educators to ask their clients and patients about their plans for sexual activity and to counsel them in ways supportive of their sexual health.

Our research had several limitations. First, we did not assess the infection status of participants, their sexual partner(s), or household members. We also did not measure whether or not a participant or anyone around them had exhibited COVID-19 symptoms in the past month. Future studies could explore the impact of infection status and duration, as well as symptom experiences, on sexual decision-making. Second, due to space limitations and relative population-level infrequency of anal behaviors,^37^ we did not ask about any changes anal sex activities. The potential of feces as a virus transmission route warrants further investigation of these behaviors. Third, we did not ask participants about their formal inclusion in a stay-at-home/shelter-in-place order and/or the extent to which they were following such an order, though we do know that most of the country was subjected to such in the month prior to the study period. This information could have implications for the structure of time available for sex, particularly in the context of other obligations like work or childcare. Finally, this survey assessed sexual behavior changes relatively early in the epidemic. We do not know how measures taken to manage COVID-19 could change behavioral practices in ways that could increase or reduce odds to adverse sexual outcomes.

## Conclusion

As the COVID-19 continues, emerging SRH focused research should continue to focus on the pandemic’s impacts on people’s sexuality. Information about types and frequency of sexual behavior are key to both aiding public health officials to disseminate accurate information and to, as well as to helping medical and sexual health education professionals proactively counsel their patients/clients. More data help achieve full understanding of transmission of the SARSCoV-2 virus through intimate contact, as well as to understand the short- and long-term influence of the COVID-19 pandemic on sexual health and well-being.

## Data Availability

Data and code underlying the findings presented in this paper are available from the first author upon request.

